# Do antibody positive healthcare workers have lower SARS-CoV-2 infection rates than antibody negative healthcare workers? Large multi-centre prospective cohort study (the SIREN study), England: June to November 2020

**DOI:** 10.1101/2021.01.13.21249642

**Authors:** V Hall, S Foulkes, A Charlett, A Atti, EJM Monk, R Simmons, E Wellington, MJ Cole, A Saei, B Oguti, K Munro, S Wallace, PD Kirwan, M Shrotri, A Vusirikala, S Rokadiya, M Kall, M Zambon, M Ramsay, T Brooks, SIREN Study Group, CS Brown, MA Chand, S Hopkins

## Abstract

**Background:** There is an urgent need to better understand whether individuals who have recovered from COVID-19 are protected from future SARS-CoV-2 infection.

**Methods:** A large multi-centre prospective cohort was recruited from publicly funded hospital staff in the UK. Participants attended regular SARS-CoV-2 PCR and antibody testing (every 2-4 weeks) and completed fortnightly questionnaires on symptoms and exposures. At enrolment, participants were assigned to either the positive cohort (antibody positive or prior PCR/antibody test positive) or negative cohort (antibody negative, not previously known to be PCR/antibody positive). Potential reinfections were clinically reviewed and classified according to case definitions (confirmed, probable, possible (subdivided by symptom-status)) depending on hierarchy of evidence. Individuals in the primary infection were excluded from this analysis if infection was confirmed by antibody only. Reinfection rates in the positive cohort were compared against new PCR positives in the negative cohort using a mixed effective multivariable logistic regression analysis.

**Findings:** Between 18 June and 09 November 2020, 44 reinfections (2 probable, 42 possible) were detected in the baseline positive cohort of 6,614 participants, collectively contributing 1,339,078 days of follow-up. This compares with 318 new PCR positive infections and 94 antibody seroconversions in the negative cohort of 14,173 participants, contributing 1,868,646 days of follow-up. The incidence density per 100,000 person days between June and November 2020 was 3.3 reinfections in the positive cohort, compared with 22.4 new PCR confirmed infections in the negative cohort. The adjusted odds ratio was 0.17 for all reinfections (95% CI 0.13-0.24) compared to PCR confirmed primary infections. The median interval between primary infection and reinfection was over 160 days.

**Interpretation:** A prior history of SARS-CoV-2 infection was associated with an 83% lower risk of infection, with median protective effect observed five months following primary infection. This is the minimum likely effect as seroconversions were not included.

**Funding:** Department of Health and Social Care and Public Health England, with contributions from the Scottish, Welsh and Northern Irish governments.

## 1. BACKGROUND

There is an urgent need to better understand whether individuals who have recovered from COVID-19 are protected from future SARS-CoV-2 infection.^1,2^ Establishing whether reinfection is typically symptomatic or asymptomatic, whether reinfected individuals are infectious to others and the expected duration of SARS-CoV-2 immunity from infection and vaccination are key components of determining the future dynamics of SARS-CoV-2 circulation.

Reinfections have been reported internationally since June 2020, although they remain uncommon.^2-21^ Large longitudinal cohort studies with regular testing are needed to understand the rates of reinfection and their implications for policy by providing systematic epidemiological, virological, immunologic and clinical data.^22^

Over 90% of individuals infected with SARS-CoV-2 develop antibodies about one week after symptoms onset, persisting for at least three months.^23,24^ High levels of neutralising antibodies targeting SARS-CoV-2 Spike protein offer considerable protection against SARS-CoV-2 reinfection, supported by data from common human coronaviruses and non-human primate models and vaccine studies.^25-29^ Whilst the exact length of immunity conferred by natural infection is still unknown, titres of neutralising antibodies against SARS-CoV-2 spike protein were detectable for at least five months after primary infection.^30^

A few studies to date have reported that individuals with SARS-CoV-2 antibodies are at lower risk of clinical reinfection than antibody negative individuals.^31-33^ However, given the relatively small size of some of these cohorts and the lack of systematic SARS-CoV-2 molecular testing, the true population impact remains unknown.

The SARS-CoV-2 Immunity and Reinfection Evaluation (SIREN) Study is a large, national, multi-centre prospective cohort study of hospital healthcare workers across the National Health Service in the United Kingdom, investigating whether the presence of antibody to SARS-CoV-2 (anti-SARS-CoV-2) is associated with a reduction in the subsequent risk of symptomatic and asymptomatic reinfection over the next year.

This paper presents an interim analysis of the primary study objective, with data collected up to 24 November 2020.

## 2. METHODS

### Study design and setting

The SIREN study is a prospective cohort study among staff working in the publicly funded hospitals (the National Health Service (NHS)) across the UK. The SIREN protocol is described elsewhere.^34^

### Participants

All healthcare workers, support staff and administrative staff working at hospital sites participating in SIREN, who could provide informed consent and anticipated remaining engaged in follow-up for 12 months were eligible to join SIREN. Participants were excluded from this analysis if they had no linked antibody or PCR data, no PCR tests after enrolment or enrolled after 9 November 2020.

### Variables

Questionnaires on symptoms and exposures were sent electronically at baseline and every two weeks (Supplementary appendix); SARS-CoV-2 antibody (using the Roche cobas® or Abbott immunoassay®) and Nucleic Acid Amplification Testing (NAAT), generally RT-PCR, was conducted at enrolment and at regular intervals (PCR every two weeks, antibody every four weeks). Testing was performed in the clinical laboratory at the site of participant enrolment, using locally validated testing platforms.

### Cohort assignment at baseline

Participants were assigned to the positive cohort if they met one of the following criteria: antibody positive on enrolment or antibody positive from prior clinical laboratory sample, with or without prior PCR positive; antibody negative on enrolment with prior antibody positive, with or without prior PCR positive; antibody negative on enrolment with a PCR positive result prior to enrolment. Participants were assigned to the negative cohort if they had a negative antibody test and no documented positive PCR test. Those in the negative cohort moved to the positive cohort 21 days following a PCR positive test result or at the time of antibody seroconversion with no positive PCR test.

### Reinfection case definitions

The SIREN case definitions for reinfections have been described elsewhere and range from confirmed to possible dependent on the strength of serological, genetic and virological evidence.^36^ A possible reinfection was defined as a participant with two PCR positive samples 90 or more days apart (based on previous national surveillance analysis)^36^ with available genomic data or an antibody positive participant with a new positive PCR at least four weeks after the first antibody positive result. A probable case additionally required supportive quantitative serological data and/or supportive viral genomic data from confirmatory samples.

We subcategorised possible reinfections by symptom status to highlight those with stronger evidence and provide comparability with definitions used elsewhere.^28,31^ Participants reporting any of cough, fever, anosmia or dysgeusia 14 days before or after their positive PCR result were defined as having ‘COVID-19 symptoms’ and ‘other potential COVID-19 symptoms’ if reporting any other recognised symptoms listed in Appendix A.^34,35^

### Data sources/measurement

Individuals consented at enrolment for all their recorded results from the Public Health England (PHE) national laboratory testing surveillance system from 1^st^ February 2020 to be included in this analysis.

### Data management and linkage

Personal identifiable information collected via the enrolment survey, completed by all SIREN participants, was used to match participants to their NHS numbers, which were obtained through the Demographic Batch Service (DBS). This information (forename, surname, date of birth and NHS number) was used to link the SIREN survey data (enrolment and follow-up survey) to results from all laboratory investigations (PCR and antibody data) held at PHE. Automated data linkage was developed and run daily to extract new test results. All SIREN data (survey and laboratory extracts) were sorted and matched in the SIREN SQL database. Data were extracted from all sources on 24 November 2020.

### Detection of potential reinfections

An SQL query was run on the SIREN database daily, to identify any participants who ‘flagged’ as a potential reinfection. This included participants who had two positive PCR tests 90 days apart or antibody positive participants with a PCR positive test four weeks after their first antibody positive date. In addition, sites were advised to report potential reinfections.

### Bias

Data were collected on potential confounders, including site and participant demographics to permit adjustment in analysis. Questionnaires were piloted and formatted to reduce misclassification bias. Recall bias was limited once enrolled by asking participants to complete surveys two weekly for exposures and symptoms. Verification that sites were using validated testing platforms and standardised criteria for reporting into SGSS was obtained during site initiation.

### Study size

Recruitment will continue until 31 March 2021, recruiting up to 100,000 participants. The study was originally powered to detect a difference in rate of infection between cohorts with a sample size of 10,000 (25% estimated to be antibody positive at baseline),cumulative incidence of 2% and immune efficacy of at least 50%.^34^ The interim analysis was conducted as the cumulative incidence in the total cohort reached 2%.

### Quantitative variables: Person time at risk

Data was censored on 24 November 2020, with the following cohorts assigned.

a. Cohort susceptible to primary infection: From first antibody negative date or first PCR positive date or seroconversion (if no PCR positive reported prior to seroconversion); or if neither of these occurred, to censor date.
b. Cohort with prior infection: the earliest date for prior infection was taken as whichever is first of the PCR positive result, onset of symptoms if there was no PCR positive, or if neither is available the first positive antibody test.

### Statistical methods

The cohort was described by their baseline cohort allocation. Participants with PCR positive results in both negative and positive cohorts, were described in more detail. Cumulative incidence, using the total number of participants in each cohort, and incidence density using the total person time at risk was calculated for both cohorts and sub-categories and plotted over time using PCR confirmation only. A mixed effects logistic regression analysis was used to estimate odd ratios (OR) to measure the association between the exposure (cohort allocation) and the binary outcome (PCR test result). The entry date used in this analysis for all participants was the earliest antibody test. All PCR tests after the entry date have been used, except PCR tests within 21 days of a positive PCR result. To account for temporal changes in the background risk of infection, all tests were allocated to the calendar week of the test date. These were categorised into nine groups; <week 31, then two-week groups up to the final category of >week 44, allowing incidence over time to vary in a stepwise constant manner. Study site was fitted as a random effect to account for the longitudinal nature of the study data, with age group, gender, ethnicity, staff group, and region fitted as non-time varying fixed effects to account for their possible confounding effect.^37^ Analysis was conducted in STATA v15.1 (College Station, TX: StataCorp LLC).

## 3. RESULTS

From 18 June to 09 November 2020, 20,787 enrolled participants, with linked data on antibody and PCR testing, were included in this analysis (figure 1). The baseline cohorts assigned 6,614 (32%) to the positive cohort and 14,173 (68%) to the negative cohort. A full description of the SIREN cohort and baseline risk factors for antibody positivity is published separately.^35^ Table 1 describes the SIREN participants by their baseline cohort assignment; in summary the cohort was predominately female (n=17,487; 84%), white (n=18,304; 88%), middle-aged (median age 45.9, interquartile range 35.8-53.6) and from clinical occupations with representation from all English regions and two-thirds of acute hospital trusts.

**Table 1:** Demographics of SIREN participants by baseline cohort allocation, participants enrolled 18 June to 09 November 2020 (n=20,787)

| Characteristics |  | Positive cohort | Negative cohort | All Participants |
| --- | --- | --- | --- | --- |
|  |  | n (%) | n (%) | n (%) |
| **Gender** |  |  |  |  |
|  | Female | 5452 (82.4) | 12035 (84.9) | 17487 (84.1) |
|  | Male | 1152 (17.4) | 2121 (15.0) | 3273 (15.7) |
|  | Other | 10 (0.2) | 17 (0.1) | 27 (0.1) |
| **Age** |  |  |  |  |
|  | Median [IQR] | 46 [35.1-54] | 45.9 [36-53.9] | 45.9 [35.8-53.6] |
|  | Range | 18.6-78.4 | 18.6-84.3 | 18.6-84.3 |
| **Ethnicity** |  |  |  |  |
|  | White | 5607 (84.8) | 12697 (89.6) | 18304 (88.1) |
|  | Mixed Race | 558 (8.4) | 780 (5.5) | 1338 (6.4) |
|  | Asian | 188 (2.8) | 226 (1.6) | 414 (2.0) |
|  | Black | 102 (1.5) | 211 (1.5) | 313 (1.5) |
|  | Chinese | 111 (1.7) | 161 (1.1) | 272 (1.3) |
|  | Other Ethnic Group | 38 (0.6) | 74 (0.5) | 112 (0.5) |
|  | Prefer not to say | 10 (0.2) | 24 (0.2) | 34 (0.2) |
| **Staff group** |  |  |  |  |
|  | Nursing/Healthcare Assistant | 2964 (44.8) | 5805 (41.0) | 8769 (42.2) |
|  | Administrative/Executive | 885 (13.4) | 2311 (16.3) | 3196 (15.4) |
|  | Doctor | 793 (12) | 1423 (10) | 2216 (10.7) |
|  | Specialist staff | 340 (5.1) | 783 (5.5) | 1123 (5.4) |
|  | Healthcare Scientist | 176 (2.7) | 548 (3.9) | 724 (3.5) |
|  | Midwife | 163 (2.5) | 379 (2.7) | 542 (2.6) |
|  | Pharmacist | 78 (1.2) | 192 (1.4) | 270 (1.3) |
|  | Estates/Porters/Security | 56 (0.8) | 103 (0.7) | 159 (0.8) |
|  | Other | 1159 (17.5) | 2629 (18.5) | 3788 (18.2) |
| **Medical conditions** |  |  |  |  |
|  | No medical conditions | 4944 (74.8) | 10590 (74.7) | 15534 (74.7) |
|  | One to two medical conditions | 1635 (24.7) | 3512 (24.8) | 5147 (24.8) |
|  | Over two medical conditions | 35 (0.5) | 71 (0.5) | 106 (0.5) |
| **Region** |  |  |  |  |
|  | South West | 1016 (15.4) | 3762 (26.5) | 4778 (23) |
|  | North West | 1074 (16.2) | 1730 (12.2) | 2804 (13.5) |
|  | London | 1031 (15.6) | 1570 (11.1) | 2601 (12.5) |
|  | South East | 748 (11.3) | 1685 (11.9) | 2433 (11.7) |
|  | East Midlands | 759 (11.5) | 1586 (11.2) | 2345 (11.3) |
|  | East of England | 553 (8.4) | 1516 (10.7) | 2069 (10) |
|  | West Midlands | 710 (10.7) | 977 (6.9) | 1687 (8.1) |
|  | Yorkshire and the Humber | 541 (8.2) | 1040 (7.3) | 1581 (7.6) |
|  | North East | 182 (2.8) | 307 (2.2) | 489 (2.4) |
| **All Participants** |  | **6614 (31.8)** | **14173 (68.2)** | **20787 (100.0)** |

**Table 2:** Characteristics of reinfections and new infections detected in SIREN participants up to 24 November 2020, stratified by case definition (n=362)

|  |  | Positive cohort |  |  | Negative cohort |
| --- | --- | --- | --- | --- | --- |
|  |  | Probable<br>n (%) | Symptomatic<br>possible<br>n (%) | All probable/<br>possible<br>n (%) | New PCR+<br>n (%) |
| **Gender** |  |  |  |  |  |
|  | Female | 2 (100) | 11 (84.6) | 36 (81.8) | 261 (82.1) |
|  | Male | 0 (0) | 2 (15.4) | 8 (18.2) | 56 (17.6) |
|  | Other | 0 (0) | 0 (0) | 0 (0) | 1 (0.3) |
| **Age** |  |  |  |  |  |
|  | Median (range) | 41.5 (37-46) | 46 (25-58) | 48.5 (23-63) | 45.3 (19-70) |
| **Antibody status at baseline** |  |  |  |  |  |
|  | Positive | 2 (100) | 12 (92.3) | 40 (90.9) | 0 (0) |
|  | Negative | 0 (0) | 1 (7.7) | 3 (6.8) | 310 (97.5) |
|  | Indeterminate/not available | 0 (0) | 0 (0) | 1 (2.3) | 8 (2.5) |
| **Reinfection PCR semi quantitative values (CT/RLU)** |  |  |  |  |  |
|  | CT range (n) | 21-24 (2) | 13-37 (5) | 13-45 (16) | - |
|  | RLU range (n) | - | 587-1193 (6) | 591-1260 (20) | - |
| **Symptom status +/-14 days reinfection PCR+** |  |  |  |  |  |
|  | COVID-19 symptoms | 1 (50) | 3 (23.1) | 4 (9.1) | 196 (61.6) |
|  | Any other symptoms | 1 (50) | 10 (76.9) | 11 (25) | 53 (16.7) |
|  | No symptoms | 0 (0) | 0 (0) | 21 (47.7) | 40 (12.6) |
|  | Not known | 0 (0) | 0 (0) | 8 (18.2) | 29 (9.1) |
| **Time interval in days – median (range); n** |  |  |  |  |  |
|  | Symptom onset first episode to reinfection PCR | 212 (197-227); 2 | 166 (90-223); 10 | 169 (90-227); 32 | - |
|  | First positive PCR to reinfection PCR | - | 155 (95-201); 7 | 162 (95-223); 21 | - |
|  | First antibody positive to reinfection PCR | 63 (62-64); 2 | 110 (35-136); 12 | 101.5 (35-174); 42* | - |
| **Total** |  | **2** | **13** | **44** | **318** |

\*One participant never antibody positive, one participant first reported as antibody positive on the same date as reinfection PCR date.

**Figure 1:**
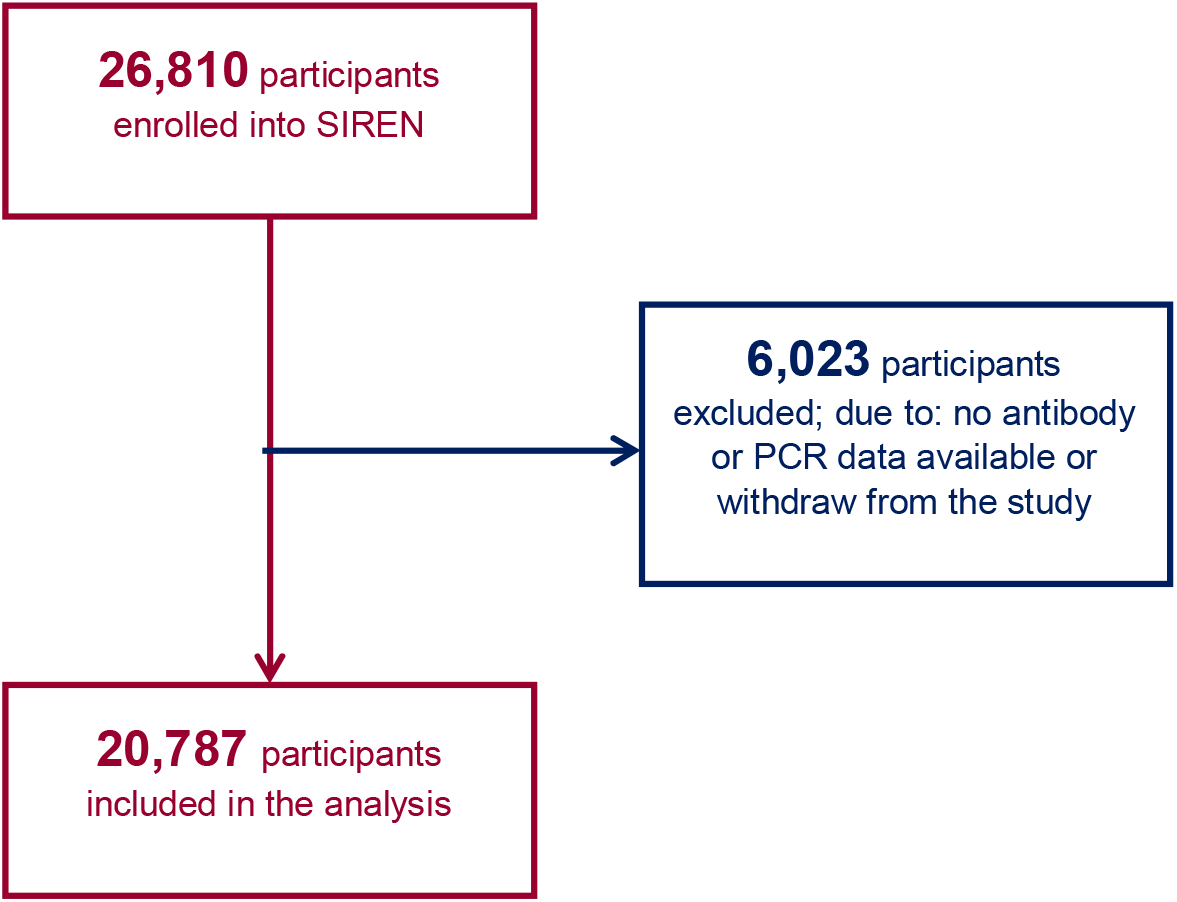
Flow diagram describing participant flow and exclusion criteria for participants enrolled in SIREN between 18 June and 09 November 2020.

The cohort had 129,189 PCR tests (17,538 before SIREN enrolment and 111,651 after enrolment) and 91,165 antibody tests (13,867 before SIREN enrolment and 77,298 after enrolment); median (and interquartile range) number of post enrolment PCR and antibody tests were 5 (3-7) 3 (2-5) respectively.

Figure 2 describes the weekly total of new PCR positives (primary infection only) in SIREN participants between March and November 2020 by baseline cohort assignment It demonstrates that PCR positivity in the positive cohort peaked in the first week of April in the negative cohort was in the last week of October 2020.

**Figure 2:**
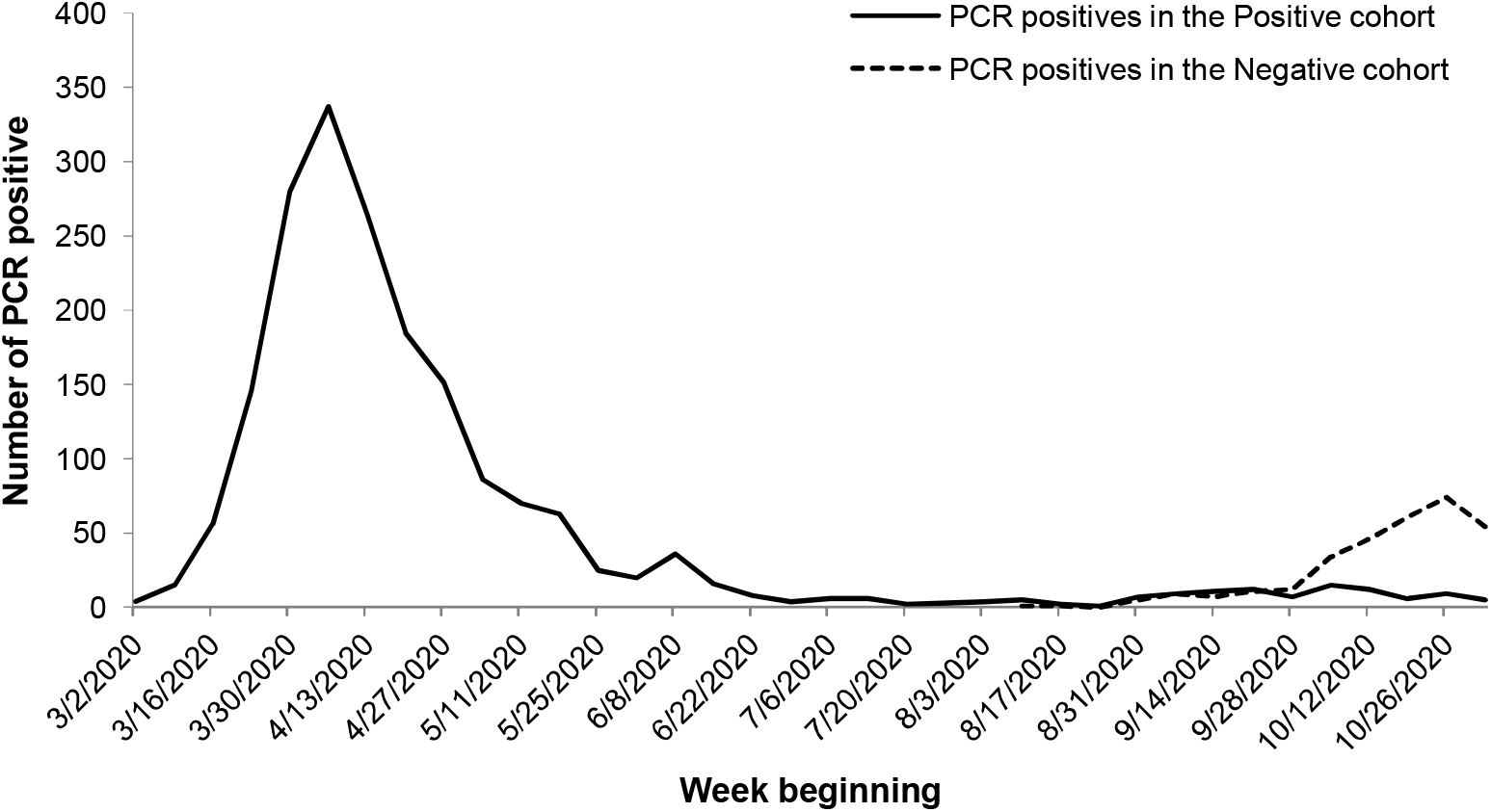
Weekly frequency of SIREN participants with a first PCR positive test result by baseline cohort assignment, March to 24 November 2020.

By 24 November 2020, 409 new infections were detected in the negative cohort: 318 were new PCR positive infections; 249 (79%) of these cases were symptomatic at infection, 196 (62%) with typical COVID-19 symptoms, and 53 (17%) with other symptoms; 40 (12%) were asymptomatic and 28 (9%) did not complete a questionnaire at the time of their symptoms; 94 were seroconversions in participants without a positive PCR test; these are not included in this interim analysis.

Forty-four reinfections were identified, 15 (34%) were symptomatic: two defined as probable (described in detail elsewhere^36)^, both symptomatic, and 42 possible; 13 symptomatic, two (23%) of whom reported typical COVID-19 symptoms. Forty (both probable and 38 possible) reinfections were antibody positive at enrolment; three had previously positive antibody tests but two were antibody negative and one indeterminate on enrolment; and one individual remained antibody negative but reported COVID-19 symptoms and a documented PCR positive status in April 2020. Twenty-one (47.7%)(50%) of these individuals had historic PCR positives from their primary infection, of whom 19 reported COVID-19 symptoms and two other symptoms within 14 days of their positive test. Fourteen (31.8%) individuals (including both probable cases) reported a history of COVID-19-like illness but did not have a PCR test due to lack of availability at the time of their primary illness; 13 (92.9%) with typical COVID-19 symptoms and one with other symptoms. Nine (20.5%) reported no history of any potential COVID-19 related symptoms.

For the 32 reinfections providing a history of COVID-19 symptoms, used as a proxy to estimate the date of their primary infection, the median interval between primary infection and reinfection beyond 90 days was 172 days (90-227) and for the 21 reinfections with a historic PCR positive test before enrolment, the median interval between the historic PCR positive date and the reinfection PCR positive date was 162 days (95-223).

Between June and November 2020, the cumulative incidence of probable, symptomatic possible and all reinfections in the positive cohort between June and November 2020 was 0.3, 2.3 and 6.7 per 1,000 participants respectively and incidence of symptomatic and all new PCR infections in the negative cohort was 17.6 and 22.4 per 1,000 participants respectively (Table 3). The incidence density per 100,000 days of follow up between June and November 2020 in the positive cohort was 3 .3 reinfections and in the negative cohort was 17.0 new PCR positive infections per 100,000 days of follow-up. Figure 3 describes the cumulative incidence of new episode PCR positive tests per cohort demonstrating the higher cumulative incidence in the negative cohort reaching 20 per 1000 compared 5 per 1000 cases in the positive cohort.

**Table 3:** Frequency of new infections and possible/probable reinfections by cohort, characterised by symptoms within 14 days (pre/post) of PCR positive date and exposures in preceding 14 days (n=362)

|  | Positive cohort |  |  |  |  |  | Negative cohort |  |  |  |  |  |
| --- | --- | --- | --- | --- | --- | --- | --- | --- | --- | --- | --- | --- |
|  | n | Denominator |  | Incidence |  |  | n | Denominator |  | Incidence |  |  |
|  |  | Total participants | Person time at risk (days) | Cumulative (per 1000) | Density (per 100,000) |  |  | Total participants | Person time at risk (days) | Cumulative (per 1000) | Density (per 100,000) |  |
| Reinfections |  |  |  |  |  |  | New PCR |  |  |  |  |  |
| Probable | 2 | 6614 | 1,339,078 | 0.3 | 0.1 |  | - | - | - | - | - | - |
| Probable AND Symptomatic possible | 15 | 6614 | 1,339,078 | 2.3 | 1.1 |  | 249 | 14,173 | 1,868,646 | 17.6 | 13.3 |  |
| All events | 44 | 6614 | 1,339,078 | 6.7 | 3.3 |  | 318 | 14,173 | 1,868,646 | 22.4 | 17.0 |  |

**Figure 3:**
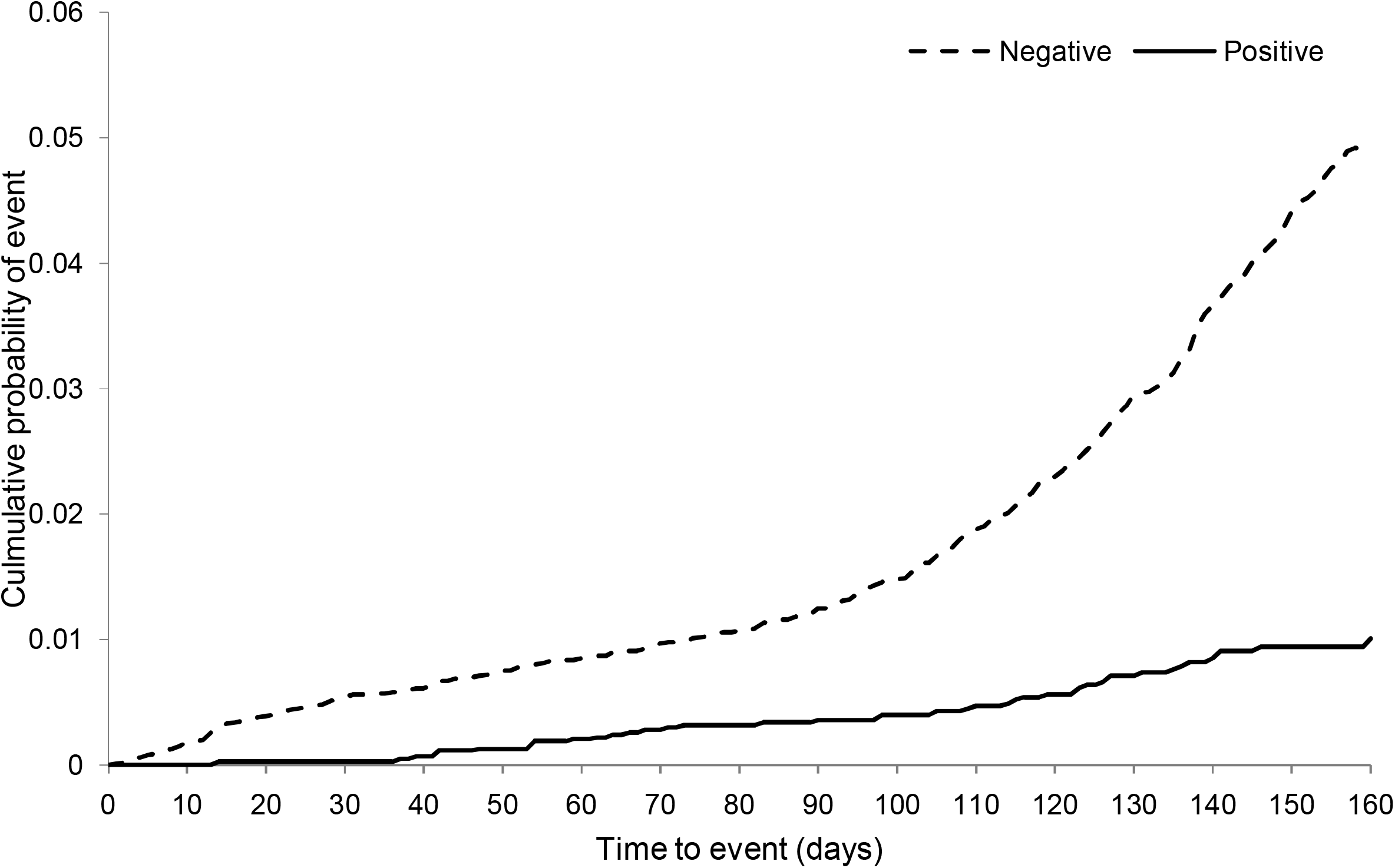
Time to PCR positive result by cohort in SIREN participants, detected up to 24 November 2020. Note: 318 PCR positive results were reported in the negative cohort; 44 PCR reinfections (probable and possible reinfections) were detected in the positive cohort during SIREN follow-up to 24 November 20202. In the positive cohort follow-up time started from date of primary PCR positive, or primary symptom onset (if no historic PCR positive and history of COVID-19 symptoms reported) or date first antibody positive. In the negative cohort follow-up started at the date of first negative antibody result. Follow-up time has been truncated at 160 days due to the size of the risk-sets becoming very small

We estimated the relative odds for reinfections in the positive cohort, with separate analyses for each reinfection definition described above, compared to new PCR positive infections in our negative cohort between SIREN enrolment and 24 November 2020 (Table 4, annex B Tables Bi.-Biii).

**Table 4:**
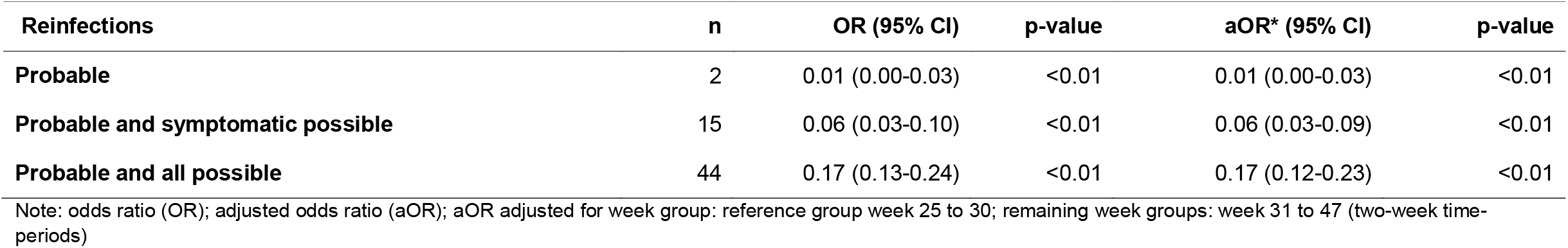
Univariable and multivariable analysis of risk of infection by cohort during SIREN follow-up, using a range of reinfection case definitions, between 18 June and 24 November 2020.

| Reinfections | n | OR (95% CI) | p-value | aOR* (95% CI) | p-value |
| --- | --- | --- | --- | --- | --- |
| Probable | 2 | 0.01 (0.00-0.03) | <0.01 | 0.01 (0.00-0.03) | <0.01 |
| Probable and symptomatic possible | 15 | 0.06 (0.03-0.10) | <0.01 | 0.06 (0.03-0.09) | <0.01 |
| Probable and all possible | 44 | 0.17 (0.13-0.24) | <0.01 | 0.17 (0.12-0.23) | <0.01 |

Note: odds ratio (OR); adjusted odds ratio (aOR); aOR adjusted for week group: reference group week 25 to 30; remaining week groups: week 31 to 47 (two-week time-periods)

Restricting reinfections to probable reinfections only, we estimated that between June and November 2020, participants in the positive cohort had 99% lower odds of probable reinfection, adjusted OR (aOR) 0.01 (95% CI 0.00-0.03). Restricting reinfections to those who were symptomatic we estimated participants in the positive cohort had 95% lower odds of reinfection, aOR 0.08 (95% CI 0.05-0.13). Using our most sensitive definition of reinfections, including all those who were possible or probable the adjusted odds ratio was 0.17 (95% CI 0.13-0.24).

The two approaches to account for temporal changes in incidence provided very similar estimates, we have opted to present results from the model with calendar time categorised. This also shows how the probability of exposure to an infectious individual has changed over time in a piecewise constant manner, increasing over time as incidence of new infections in the population increased in September and October 2020.

## 4. DISCUSSION

We have presented the interim findings after five months of follow-up from the SIREN study, a unique large-scale multi-centre prospective cohort study of healthcare staff undergoing frequent asymptomatic testing, powered to detect and characterise reinfections and estimate the protective effect of SARS-CoV-2 antibodies.

We have detected two probable reinfections (both symptomatic with high viral loads, genome sequencing demonstrating phylogenetic relatedness to concurrently circulating strains, and a boosted antibody response), which have been characterised and reported separately, ^36^ and 42 possible reinfections in our positive cohort. This compares with 318 new PCR positive infections, 249 of whom were symptomatic, 78% with typical COVID-19 symptoms, in our negative cohort. Using a symptomatic case definition aligned with positive PCR results, previous infection reduced the odds of infection by at least 90% (aOR 0.06 with 95%CI of 0.03 to 0.09) and even when we included all possible and probable reinfections reduced the odds of reinfection by at least 75% [aOR 0.17 (95% CI 0.13-0.24)]

We believe this is the minimum likely impact as the curve in the positive cohort was gradual throughout, indicating some of these potential reinfections were likely residual RNA detection at low population prevalence rather than true reinfections. In the negative cohort the gradient was gradual up to around day 100 and has then accelerated, broadly coinciding with the period when community prevalence increased rapidly.^38^ In addition, we did not include 94 seroconversions in the negative cohort, as these seroconversions were not detected by PCR and we cannot currently say whether a similar rate of undetected infections occurred in the positive cohort. None of the reinfections we have identified are confirmed by our stringent case definitions; most we only consider possible and are undergoing further serological investigation. Investigations have been restricted by the limited availability of data and samples from historic infections, with most swabs discarded without sequencing, preventing the genomic comparison between infection episodes required to confirm a reinfection. This highlights the importance of SIREN, through which we are ensuring the data collection and characterisation of new infections, to build a stronger base to investigate and confirm future reinfections. Our use of hierarchical case definitions identifies cases with stronger evidence, and allows us present the range of potential reinfection scenarios.

Another limitation is measurement error capturing the primary infection onset date for positive cohort participants without a PCR positive test associated with their primary episode. This introduces imprecision into both our person time at risk, and consequently reinfection rates, and our estimated intervals between primary infection episodes and reinfections. For those who were symptomatic in their primary episode we have used their self-reported COVID-19 symptom onset date as a proxy, which may be subject to recall bias. We have introduced validation rules here to reduce this, excluding onset dates before March 2020. However, for participants with asymptomatic or non-COVID-19 symptomatic primary infections, we are reliant on using their first antibody positive date. We are therefore not capturing all the time they were susceptible to reinfection, reducing our overall follow-up time for this cohort, and thus inflating our reinfection rates and reducing our intervals between infection episodes.

As the cohort assignment has been determined by testing at SIREN sites, which use a range of testing platforms and assays, there is the possibility of misclassification bias. We have included participants in the positive cohort who had a prior positive PCR test, irrespective of their antibody status. Some of those PCR results, especially early in the epidemic, may have been false positives or laboratory contamination episodes, particularly given Ct/RLU values are not available. We aim to retest all baseline serum samples within PHE, using both S and N target assays in order to give each participant a validated quantitative baseline antibody result. This will inform future analyses and may lead to changes to the cohort assignment presented.

Finally, this interval analysis is presented prior to the widespread emergence and spread of the B1.1.7 lineage (VOC202012/01) with multiple non-synonymous spike mutations including N501Y; the impact of this lineage on future protection remains undetermined and will be evaluated.

Our results are consistent with the findings from other smaller studies of decreased incidence of PCR positivity in antibody positive individuals.^31,39^ Another prospective cohort of healthcare workers recently reported the incidence of new positive PCR-confirmed infections to be lower among seropositive than seronegative participants (n=3/1,246 vs. n=165/11,052, an incidence density of 2.1 and 8.6/100,000 days at risk respectively).^31^ None of the three potential reinfections were symptomatic.

The recent SARS-CoV-2 vaccination trials have typically investigated protection from symptomatic infection. The ChAdOx1 trial reported protection against symptomatic infection of between 70.4% and 90%, and the BNT162b2 vaccine phase 3 results report 95% protection over two and three months of follow-up respectively.^28,29^ Our findings, after a longer period of follow-up, of 94% lower odds of symptomatic infection, demonstrate equivalent, or higher protection from natural infection, both for symptomatic and asymptomatic infection.

After five months of follow-up, this large observational study has found that prior SARS-CoV-2 infection protects most individuals against reinfection for at least five months. We have identified and investigated more potential reinfections than reported in the global literature to date, supporting the value of large prospective cohort studies such as SIREN. This study supports the hypothesis that primary infection with SARS-CoV-2 provides a high degree of immunity to repeat infection in the short to medium term; with similar levels of prevention of symptomatic infection as current licensed vaccines for working age adults. Primary infection also reduces the risk of asymptomatic infection and thus onward transmission; this is particularly important in the healthcare was considered as a potential driver for ongoing community transmission in Wave 1 in the UK.^40^ This increases the likelihood that this may also be attainable by vaccine induced immunity. Further detailed studies on the longevity of antibody responses, assessment of reinfection rates under the challenge of the new lineage, and the impact of all COVID-19 vaccines introduced in the UK are underway in this cohort.

## Data Availability

Data will be available to trusted researchers once the final study data is released.

## Trial Registration

IRAS ID 284460, HRA and Health and Care Research Wales approval granted 22 May 2020. Trial ID: ISRCTN11041050.

## Acknowledgements

thanks to all staff supporting study delivery at participating sites and to all participants for their ongoing commitment and contributions to this study. Thank you to all staff undertaking COVID-19 testing within PHE Reference Microbiology.

## Appendix A Symptom list in questionnaire

Cough, Fever, Anosmia, Dysgeusia, Sore throat, runny nose, headache, muscle aches, fatigue, diarrhoea, vomiting, itchy red patches.

## Appendix B

**Table Bi:** Multivariable analysis of risk of infection by cohort during SIREN follow-up, probable reinfections, between 18 June and 24 November 2020 (n=2)

| Characteristics | OR (95% CI) | p-value |
| --- | --- | --- |
| Positive | 0.01 (0.00-0.03) | <0.01 |
| **Week group** |  |  |
| 25 to 30 | - | - |
| 31 to 32 | 0.12 (0.04-0.32) | <0.01 |
| 33 to 34 | 0.14 (0.06-0.32) | <0.01 |
| 35 to 36 | 0.27 (0.15-0.49) | <0.01 |
| 37 to 38 | 0.48 (0.31-0.75) | <0.01 |
| 39 to 40 | 0.40 (0.25-0.62) | <0.01 |
| 41 to 42 | 0.99 (0.68-1.45) | 0.97 |
| 43 to 44 | 1.05 (0.73-1.52) | 0.79 |
| 45 to 47 | 0.20 (0.12-0.32) | 0.00 |
| **Age group** |  |  |
| 18 to 24 | - | - |
| 25 to 34 | 0.70 (0.44-1.11) | 0.13 |
| 35 to 44 | 0.46 (0.29-0.73) | <0.01 |
| 45 to 54 | 0.64 (0.41-1.00) | 0.05 |
| 55 to 64 | 0.48 (0.30-0.78) | <0.01 |
| Over 65 | 0.25 (0.07-0.83) | 0.02 |
| **Gender** |  |  |
| Male | - | - |
| Female | 0.83 (0.63-1.09) | 0.18 |
| Other | 1.40 (0.18-10.60) | 0.74 |
| **Ethnicity** |  |  |
| White | - | - |
| BAME | 1.51 (1.12-2.04) | 0.01 |
| **Staff group** |  |  |
| Nursing/Healthcare Assistant | - | - |
| Doctor | 0.66 (0.45-0.96) | 0.03 |
| Midwife | 0.83 (0.45-1.54) | 0.56 |
| Administrative/Executive | 0.75 (0.55-1.01) | 0.06 |
| Specialist staff | 0.79 (0.50-1.24) | 0.30 |
| Estates/Porters/Security | 2.08 (0.95-4.55) | 0.07 |
| Pharmacist | 0.89 (0.41-1.93) | 0.76 |
| Healthcare Scientist | 0.48 (0.26-0.89) | 0.02 |
| Other | 0.73 (0.56-0.96) | 0.03 |
| **Region** |  |  |
| East Midlands | - | - |
| East of England | 0.52 (0.25-1.07) | 0.07 |
| London | 0.34 (0.17-0.68) | <0.01 |
| North East | 1.98 (0.75-5.24) | 0.17 |
| North West | 1.53 (0.86-2.74) | 0.15 |
| South East | 0.58 (0.31-1.09) | 0.09 |
| South West | 0.32 (0.17-0.59) | <0.01 |
| West Midlands | 1.64 (0.87-3.10) | 0.13 |
| Yorkshire and the Humber | 1.77 (0.93-3.36) | 0.08 |

**Table Bii:**
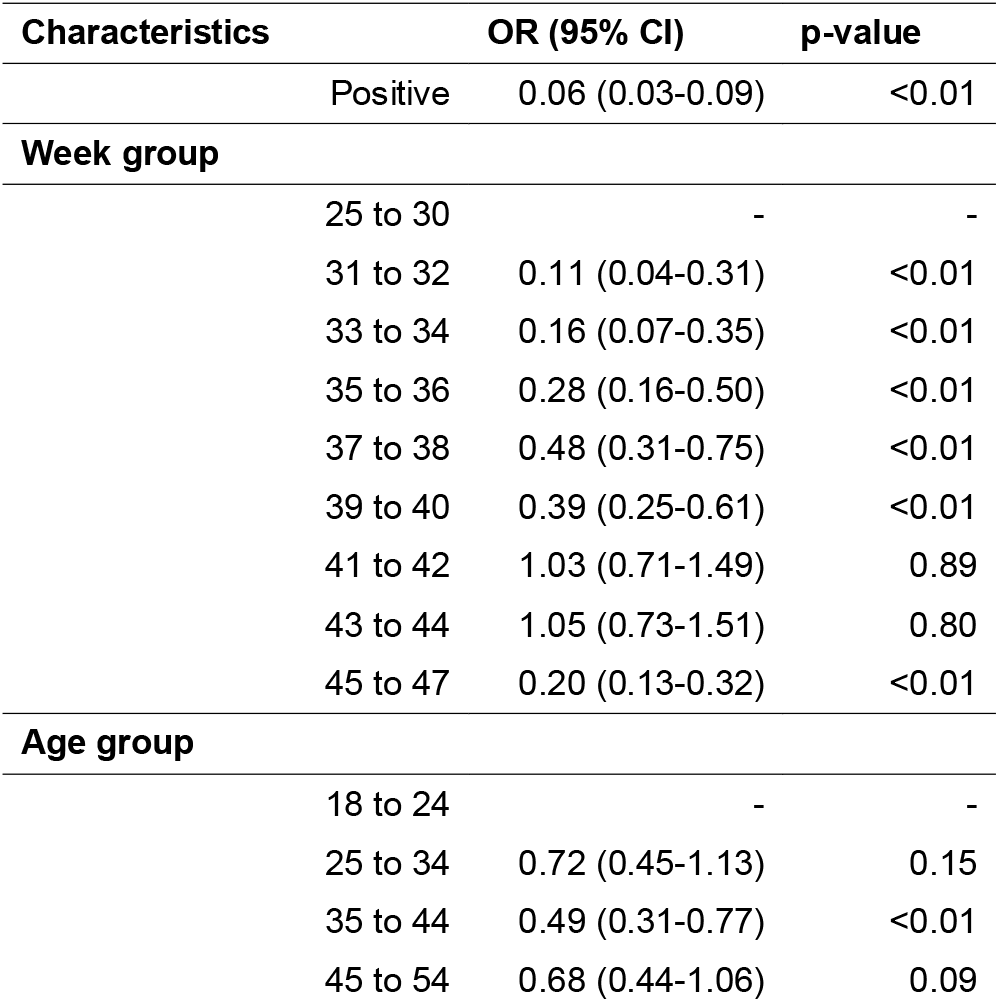
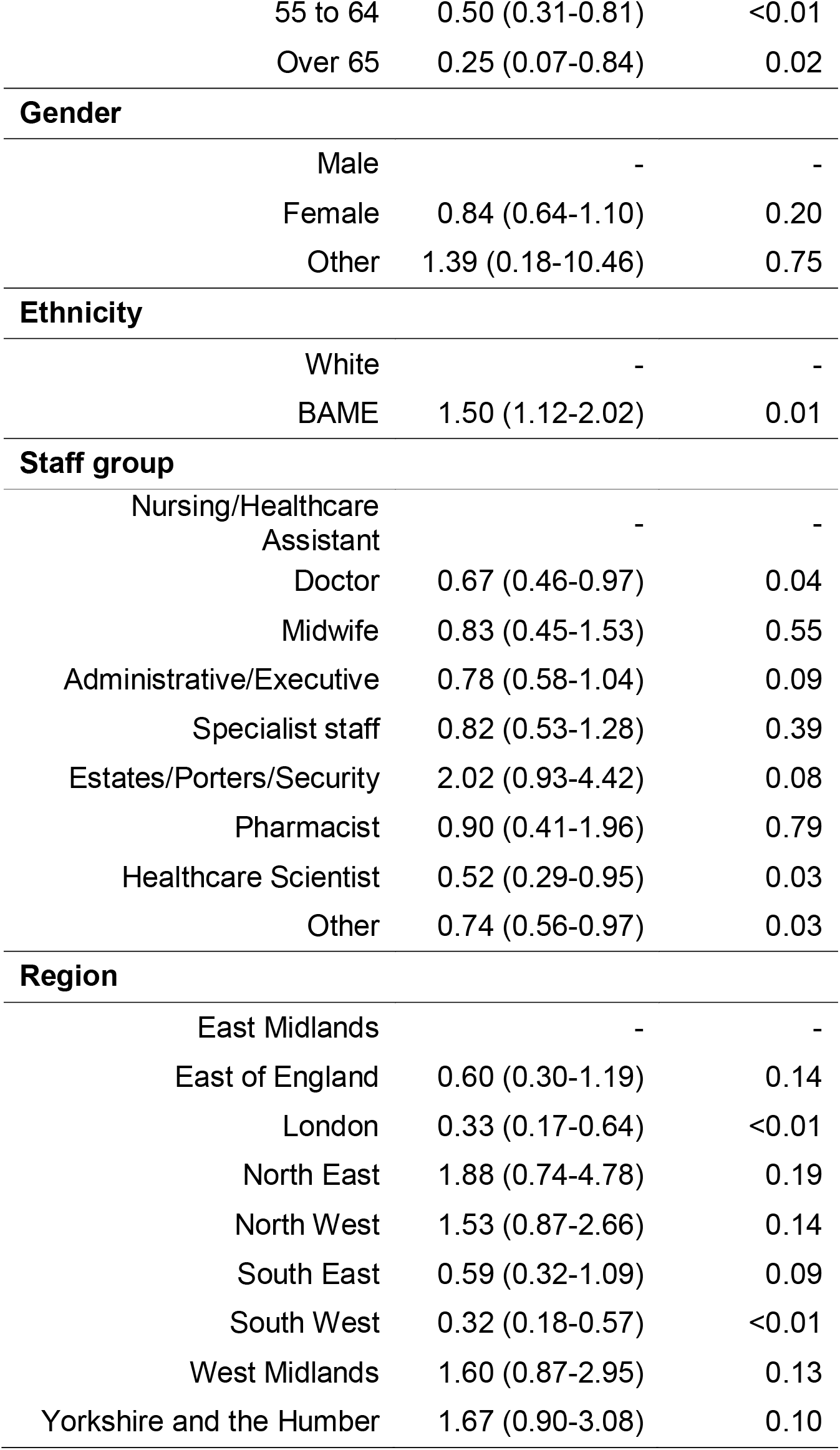
Multivariable analysis of risk of infection by cohort during SIREN follow-up, probable and symptomatic possible reinfections, between 18 June and 24 November 2020 (n=15)

| Characteristics | OR (95% CI) | p-value |
| --- | --- | --- |
| Positive | 0.06 (0.03-0.09) | <0.01 |
| **Week group** |  |  |
| 25 to 30 | - | - |
| 31 to 32 | 0.11 (0.04-0.31) | <0.01 |
| 33 to 34 | 0.16 (0.07-0.35) | <0.01 |
| 35 to 36 | 0.28 (0.16-0.50) | <0.01 |
| 37 to 38 | 0.48 (0.31-0.75) | <0.01 |
| 39 to 40 | 0.39 (0.25-0.61) | <0.01 |
| 41 to 42 | 1.03 (0.71-1.49) | 0.89 |
| 43 to 44 | 1.05 (0.73-1.51) | 0.80 |
| 45 to 47 | 0.20 (0.13-0.32) | <0.01 |
| **Age group** |  |  |
| 18 to 24 | - | - |
| 25 to 34 | 0.72 (0.45-1.13) | 0.15 |
| 35 to 44 | 0.49 (0.31-0.77) | <0.01 |
| 45 to 54 | 0.68 (0.44-1.06) | 0.09 |

FINAL MANUSCRIPT
|  |  |  |  |
| --- | --- | --- | --- |
|  | 55 to 64 | 0.50 (0.31-0.81) | <0.01 |
|  | Over 65 | 0.25 (0.07-0.84) | 0.02 |
| **Gender** |  |  |  |
|  | Male | - | - |
|  | Female | 0.84 (0.64-1.10) | 0.20 |
|  | Other | 1.39 (0.18-10.46) | 0.75 |
| **Ethnicity** |  |  |  |
|  | White | - | - |
|  | BAME | 1.50 (1.12-2.02) | 0.01 |
| **Staff group** |  |  |  |
|  | Nursing/Healthcare Assistant | - | - |
|  | Doctor | 0.67 (0.46-0.97) | 0.04 |
|  | Midwife | 0.83 (0.45-1.53) | 0.55 |
|  | Administrative/Executive | 0.78 (0.58-1.04) | 0.09 |
|  | Specialist staff | 0.82 (0.53-1.28) | 0.39 |
|  | Estates/Porters/Security | 2.02 (0.93-4.42) | 0.08 |
|  | Pharmacist | 0.90 (0.41-1.96) | 0.79 |
|  | Healthcare Scientist | 0.52 (0.29-0.95) | 0.03 |
|  | Other | 0.74 (0.56-0.97) | 0.03 |
| **Region** |  |  |  |
|  | East Midlands | - | - |
|  | East of England | 0.60 (0.30-1.19) | 0.14 |
|  | London | 0.33 (0.17-0.64) | <0.01 |
|  | North East | 1.88 (0.74-4.78) | 0.19 |
|  | North West | 1.53 (0.87-2.66) | 0.14 |
|  | South East | 0.59 (0.32-1.09) | 0.09 |
|  | South West | 0.32 (0.18-0.57) | <0.01 |
|  | West Midlands | 1.60 (0.87-2.95) | 0.13 |
|  | Yorkshire and the Humber | 1.67 (0.90-3.08) | 0.10 |

**Table Biii:**
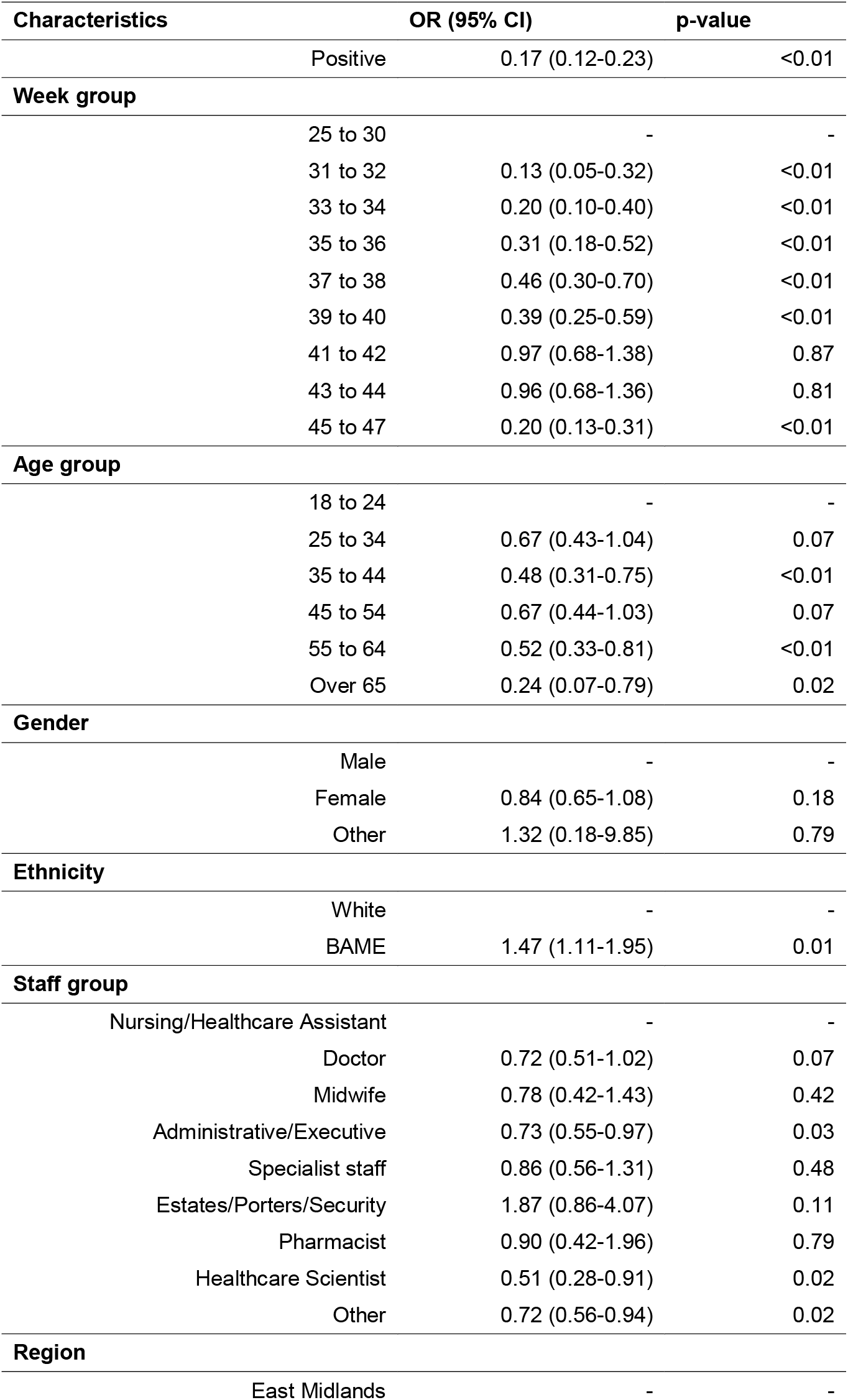
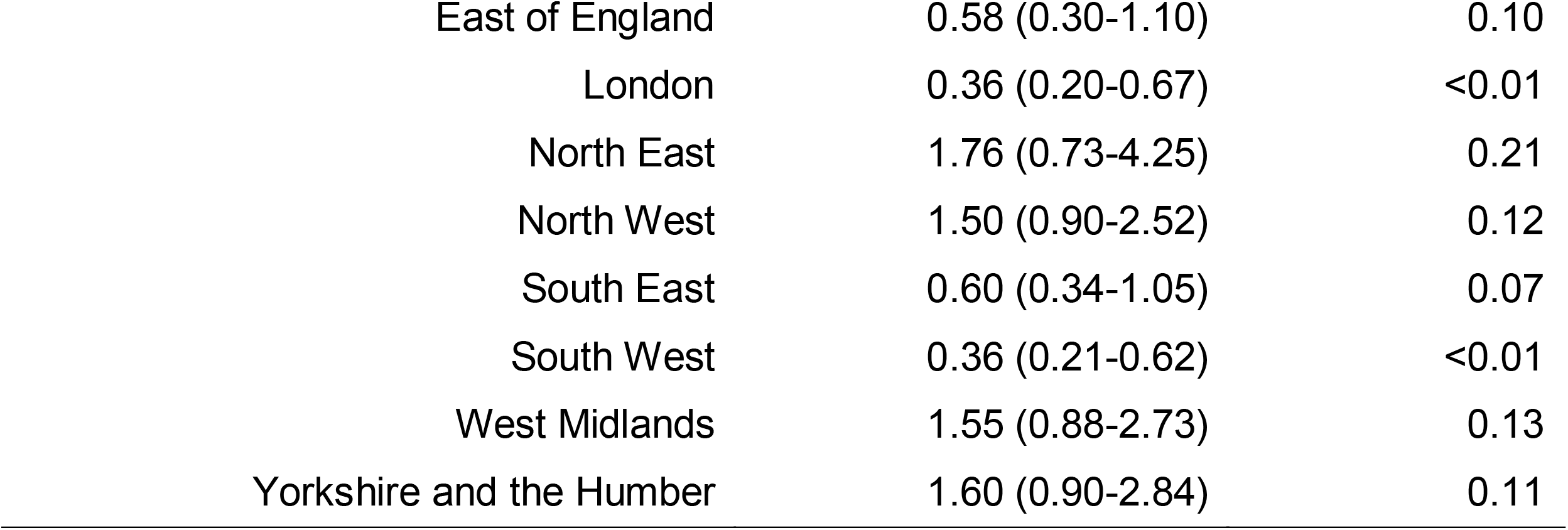
Multivariable analysis of risk of infection by cohort during SIREN follow-up, probable and all possible reinfections, between 18 June and 24 November 2020 (n=44)

**^∞^The SIREN study group**

**Public Health England**: Andrews N, A Atti, H Aziz, T Brooks, CS Brown, C Carr, MA Chand, A Charlett, H Crawford, M Cole, J Conneely, S DArcangelo, J Ellis, S Evans, S Foulkes, N Gillson, R Gopal, V Hall, P Harrington, S Hopkins, J Hewson, K Hoschler, D Ironmonger, J Islam, M Kall, I Karagiannis, J Khawam, P Kirwan, R Kyffin. A Lackenby, M Lattimore, E Linley, J Lopez-Bernal, L Mabey, R McGregor, S Miah, E Monk, K Munro, Z Naheed, A Nissr, AM O’Connell, B Oguti, S Organ, J Osbourne, A Otter, M Patel, S Platt, D Pople, K Potts, M Ramsay, J Robotham, S Rokadiya, C Rowe, A Saei, G Sebbage, A Semper, M Shrotri, R Simmons, A Soriano, P Staves, S Taylor, A Taylor Kerr, A Tengbe, S Tonge, Vusirikala A, S Wallace E Wellington & M Zambon

**Glasgow Caledonian University**: L Price (and **Public Health Scotland**), L Haahr, S Stewart.

**Health and Social Care Agency Northern Ireland**: D Corrigan, M Sartaj, L Cromey, S Campbell, K Braithwaite.

**Public Health Wales**: ED Lacey, G Stevens, L Partridge. **Health and Social Care Research Wales**: C Norman, Y Ellis, H Hodgson.

**<u>Participating SIREN sites:</u>** Site Principal and co-investigators

Alder Hey Children’s NHS Foundation Trust: S Mcwilliam, B Larru

Ashford and St Peter’s Hospitals NHS Foundation Trust: S Winchester

Basildon and Thurrock University Hospitals NHS Foundation Trust: Dr A Pai, P Cieciwa

Belfast Health & Social Care Trust: C Loughrey, A Watt

Birmingham Community Healthcare NHS Foundation Trust: F Adair, A Hawkins

Black Country Healthcare NHS Foundation Trust: A Grant, R Temple-Purcell

Blackpool Teaching Hospitals NHS Foundation Trust: J Howard, N Slawson

Bolton NHS Foundation Trust: Dr C Subudhi

Brighton and Sussex University Hospitals NHS Trust: Dr B S Davies, Dr A Bexley

Buckinghamshire Healthcare NHS Trust: N Wong, R Penn

Calderdale and Huddersfield NHS Foundation Trust: Dr G Boyd, Dr A Rajgopal

Central and North West London NHS Foundation Trust: Dr A Arenas-Pinto, R Matthews

Chesterfield Royal Hospital NHS Foundation Trust: A Whileman

Cornwall Partnership NHS Foundation Trust: Dr R Laugharne, J Ledger

Countess of Chester Hospital NHS Foundation Trust: Dr T Barnes, C Jones

Dartford and Gravesham NHS Trust: Dr N Chitalia, D Botes

Derbyshire Healthcare NHS Foundation Trust: G Harrison, S Akhtar

Devon Partnership NHS Trust: S Horne, N Walker

Doncaster and Bassetlaw Teaching Hospitals NHS Foundation Trust: K Agwuh, V Maxwell

Dorset County Hospital NHS Foundation Trust: Dr J Graves, S Williams

Dorset Healthcare University NHS Foundation Trust: Research & Development and Occupational Health departments

East Suffolk and North Essex NHS Foundation Trust: P Ridley, A O’Kelly

East Sussex Healthcare NHS Trust: Dr A Cowley

Epsom and St Helier University Hospitals NHS Trust: H Johnstone, P Swift Frimley Health NHS Foundation Trust: M Meda, J Democratis

George Eliot Hospital NHS Trust: C Callens

Gloucestershire Hospitals NHS Foundation Trust: S Hams, S Beazer

Golden Jubilee National Hospital: V Irvine

Great Western Hospitals NHS Foundation Trust: C Forsyth, B Chandrasekaran

Hampshire Hospitals NHS Foundation Trust: Dr C Thomas, J Radmore

Hounslow and Richmond Community Healthcare NHS Trust: S Roberts, K Brown

Hull University Teaching Hospitals NHS Trust: K Gajee, P Burns

Imperial College Healthcare NHS Trust: F Sanderson, T M Byrne Isle of Wight NHS Trust: Dr E Macnaughton, S Knight

James Paget University Hospitals NHS Foundation Trust: Prof B J L Burton, H Smith

King’s College Hospital NHS Foundation Trust: R Chaudhuri

Lancashire & South Cumbria NHS Foundation Trust: Dr R Shorten, K Hollinshead

Lancashire Teaching Hospitals NHS Foundation Trust: R J Shorten, H Cross

Leeds Teaching Hospitals NHS Trust: J Murira, C Favager

Leicestershire Partnership NHS Trust: Dr S Hamer, Dr S Baillon

Lewisham and Greenwich NHS Trust: Dr J Russell, K Gantert

Lincolnshire Partnership NHS Foundation Trust: Dr A Dave, D Brennan

Liverpool University Hospitals NHS Foundation Trust: Dr A Chawla, F Westell

London North West University Healthcare NHS Trust: Dr D Adeboyeku, Dr Papineni

Maidstone and Tunbridge Wells NHS Trust: C Pegg, M Williams

Manchester University NHS Foundation Trust: Dr M Mirfenderesky, J Price

Mid Cheshire Hospitals NHS Foundation Trust: C Gabriel, K Pagget

Mid Essex Hospital Services NHS Trust: G Maloney, P Cieciwa

Mid Yorkshire Hospitals NHS Trust: Dr J Ashcroft, I Del Rosario

Moorfields Eye Hospital NHS Foundation Trust: Dr. R Crosby-Nwaobi, C Reeks

NHS Fife: S Fowler

NHS Forth Valley: Dr Spears NHS Grampian: A Milne

NHS Greater Glasgow And Clyde: J Anderson

NHS Lothian: S Donaldson, K Templeton

Norfolk and Norwich University Hospitals NHS Foundation Trust: Dr N Elumogo, L Coke

North Cumbria Integrated Care NHS Foundation Trust: J Elliott, D Padgett

North Middlesex University Hospital NHS Trust: M Mirfenderesky, J Price

North West Anglia NHS Foundation Trust: I Sinanovic, S Joyce

Northern Devon Healthcare NHS Trust: Dr T Lewis, M Howard

Northern Lincolnshire and Goole NHS Foundation Trust: Dr P Cowling, D Potoczna

Nottingham University Hospitals NHS Trust: S Brand

Poole Hospital NHS Foundation Trust: Dr L Sheridan, B Wadams

Portsmouth Hospitals NHS Trust: A Lloyd, J Mouland

Queen Victoria Hospital NHS Foundation Trust: Dr J Giles, G Pottinger

Royal Berkshire NHS Foundation Trust: H Coles, M Joseph

Royal Bournemouth and Christchurch Hospitals NHS Foundation Trust: Dr M Lee, S Orr

Royal Cornwall Hospitals NHS Trust: H Chenoweth

Royal Devon and Exeter NHS Foundation Trust: C Auckland, R Lear

Royal Free London NHS Foundation Trust: Dr A Rodger, Dr T Mahungu

Royal National Orthopaedic Hospital NHS Trust: K Penny-Thomas

Royal Papworth Hospital NHS Foundation Trust: Dr. S Pai, J Zamikula

Royal Surrey County Hospital NHS Foundation Trust: E Smith, S Stone

Royal United Hospitals Bath NHS Foundation Trust: E Boldock, D Howcroft

Salisbury NHS Foundation Trust: C Thompson

Sandwell and West Birmingham Hospitals NHS Trust: Dr M Aga, P Domingos

Sheffield Children’s NHS Foundation Trust: Dr C Kerrison, S Gormley

Sheffield Teaching Hospitals NHS Foundation Trust: L Marsh, S Tazzyman

Sherwood Forest Hospitals NHS Foundation Trust: S Ambalkar, L Allsop

Shrewsbury and Telford Hospital NHS Trust: S Jose, M Beekes

Shropshire Community Health NHS Trust: J Tomlinson

Solent NHS Trust: C Price, A Jones

Somerset NHS Foundation Trust: Dr J Pepperell, M Schultz

Southend University Hospital NHS Foundation Trust: Dr J Day

Southern Health & Social Care Trust: A Boulos

Southern Health NHS Foundation Trust: E Defever, D McCracken

Southport and Ormskirk Hospital NHS Trust: Dr K Gray, K Brown

St George’s University Hospitals NHS Foundation Trust: A Houston, T Planche

St Helens and Knowsley Teaching Hospitals NHS Trust: Dr M Z QAZZAFI

Stockport NHS Foundation Trust: J Marrs, S Bennett

Surrey and Sussex Healthcare NHS Trust: Dr K Nimako, Dr B Stewart

The Clatterbridge Cancer Centre NHS Foundation Trust: Dr S Khanduri, Prof N Kalakonda

The Dudley Group NHS Foundation Trust: Dr A Ashby

The Hillingdon Hospitals NHS Foundation Trust: N Mahabir, M Holden

The Newcastle Upon Tyne Hospitals NHS Foundation Trust: B Payne, J Harwood

The Princess Alexandra Hospital NHS Trust: K Court, N Staines

The Robert Jones and Agnes Hunt Orthopaedic Hospital NHS Foundation Trust: Dr R Longfellow

The Royal Wolverhampton NHS Trust: M E Green, L E Hughes

Torbay and South Devon NHS Foundation Trust: Dr M Halkes, P Mercer

United Lincolnshire Hospitals NHS Trust: A Roebuck, Research Team LCRF

University Hospital Southampton NHS Foundation Trust: Dr E Wilson-Davies

University Hospitals Bristol and Weston NHS Foundation Trust: R Lazarus, L Gallego

University Hospitals Coventry and Warwickshire NHS Trust: L Berry, N Aldridge

University Hospitals of Derby and Burton NHS Foundation Trust: Prof F Game, Prof T Reynolds

University Hospitals of Leicester NHS Trust: C Holmes, M Wiselka

University Hospitals of Morecambe Bay NHS Foundation Trust: A Higham

University Hospitals of North Midlands NHS Trust: C Duff, M Booth

University Hospitals Plymouth NHS Trust: H Jory, J Alderton

Walsall Healthcare NHS Trust: E Virgilio

Warrington and Halton Teaching Hospitals NHS Foundation Trust: Dr M Z Qazzafi, Dr T Chin

West Suffolk NHS Foundation Trust: Dr A M Moody, Dr R Tilley

Western Health & Social Care Trust: T Donaghy

Western Sussex Hospitals NHS Foundation Trust: R Sierra, K Shipman

Whittington Health NHS Trust: N Jones, G Mills

Wirral University Teaching Hospital NHS Foundation Trust: Dr D Harvey, Y W J Huang

Wye Valley NHS Trust: Dr L Robinson, J Birch

Yeovil District Hospital NHS Foundation Trust: Dr A Broadley, S Board

York Teaching Hospital NHS Foundation Trust: N Todd, C Laven

**<u>SIREN Associated studies</u>**

Oxford University Hospital study: DW Eyre, Big Data Institute, Nuffield Department of Population Health, University of Oxford, K Jeffery, Oxford University Hospitals NHS Foundation Trust,

Protective Immunity from T cells to Covid-19 in Health workers (PITCH): S Dunachie, P Klenerman, L Turtle, C Duncan.

The Humoral Immune Correlates for COVID-19 (HICC) consortium: JL Heeney, H Baxendale, W Schwaeble

